# Executive dysfunction following SARS-CoV-2 infection: A cross-sectional examination in a population-representative sample

**DOI:** 10.1101/2022.01.01.22268614

**Authors:** Peter A. Hall, Gang Meng, Anna Hudson, Mohammad N. Sakib, Sara C. Hitchman, James McKillop, Warren K. Bickel, Geoffrey T. Fong

## Abstract

**Background:** Prior studies have documented reliable associations between SARS-CoV-2 infection and adverse cognitive impact in older adults. The current study sought to determine whether SARS-CoV-2 infection and COVID-19 symptom severity are associated with cognitive dysfunction among young adults and middled-aged adults in the general population.

**Method:** The Canadian COVID-19 Experiences Project (CCEP) survey involves 1,958 adults with equal representation of vaccinated and vaccine hesitant adults between the ages of 18 and 54 years. The sample comprised 1,958 adults with a mean age of 37 years (*SD*=10.4); 60.8% were female. The primary outcome was symptoms of cognitive dysfunction assessed via an abbreviated form of the Barkley Deficits in Executive Functioning Scale (BDEFS) and performance on a validated decision-making task.

**Results:** Young and middle-aged adults with a positive SARS-CoV-2 infection history reported a significantly higher number of symptoms of executive dysfunction (*M*_*adj*_=1.89, *SE*=0.08, *CI*: 1.74, 2.04; *n*=175) than their non-infected counterparts (*M*_*adj*_=1.63, *SE*=0.08, *CI*: 1.47,1.80; *n*=1,599; β=0.26, *p*=.001). Among those infected, there was a dose-response relationship between COVID-19 symptom severity and level of executive dysfunction, with moderate (β=0.23, *CI*: 0.003-0.46) and very/extremely severe (β= 0.69, *CI*: 0.22-1.16) COVID-19 symptoms being associated with significantly greater dysfunction, compared to asymptomatic. These effects remained reliable and of similar magnitude after controlling for age, sex, vaccination status, income, and geographic region, and after removal of those who had been intubated during hospitalization. Similar effects were found for the decision-making task.

**Conclusions:** Positive SARS-CoV-2 infection history and COVID-19 symptom severity are associated with executive dysfunction among young and middle-aged adults with no history of medically induced coma. These findings are evident on self-reported and task-related indicators of cognitive function.

## Introduction

Cognitive dysfunction is one of the potential adverse consequences of SARS-CoV-2 infection, and this risk may extend well below the age margins for increased mortality risk. It is understood that SARS-CoV-2 could impact the brain through a number of non-exclusive, indirect mechanisms including hypoxia, thrombosis, coagulopathy, cytokine storm, and megakaryocyte invasion.^1–6^ Studies of predominantly older, hospitalized patients have revealed cognitive deficits in the areas of memory, spatial navigation, attention, short-term memory, and executive function.^5–7^ Further, the cognitive impairments following SARS-Cov-2 infection may persist after the acute phase of infection,^5^ a phenomenon known as “long covid”.^8,9^

Several studies have reported reliable evidence of cognitive dysfunction among those previously infected with SARS-CoV-2.^7,10–13^ However, some of these studies are limited by non-representative samples and lack of comparison to non-infected controls in the general population. Examination of a population-based sample including asymptomatic and minimally symptomatic individuals, coupled with a control sample of non-infected individuals from the same population facilitates quantification of the reliability and magnitude of SARS-CoV-2 infection impacts on cognition, if they do indeed exist. Beyond the above, relatively little is known about the extent to which cognitive deficits are predicted by age or sex, as demographic moderators. The extent to which SARS-CoV-2 adversely impacts cognitive function among younger and middle-aged adults is relatively unknown. Of particular interest are the executive functions, which are especially susceptible to environmental and systemic insult.

Executive functions are partially supported by the lateral prefrontal cortex, as well as the medial orbitofrontal cortex (mOFC). The mOFC is of particular interest, being the brain subregion most anatomically close to the hypothesized point of SARS-CoV-2 neuroinvasion. Decision-making processes supported by the OFC can be best assessed using decision-making paradigms with heavy temporal and evaluative demands, such as a delay discounting task.^14–17^ Delay discounting is a neurobehavioral process reflecting the extent to which future rewards are devalued based on their delay in time^18^ and summarized relative balance between the prefrontal cortices and the limbic systems.^14^ Greater delay discounting is reflected in the tendency to choose a lower value option that is immediately available over a higher value option that is delayed in time.

Prior studies have shown that damage to the mOFC is associated with increased delay discounting.^16,17^ Impulsive choice of rewards is mediated by dopaminergic activity within the mOFC,^19^ in contrast with choices to avoid punishment, which are mediated by the lateral OFC.^20^ The most anterior aspect of the mOFC has further been proposed as the subregion most clearly involved in processing of abstract rewards (e.g., money), in contrast with the posterior mOFC, which is involved in computation of basic rewards (e.g., food, physical pleasure).^20^ Importantly, the anterior mOFC is located immediately superior to the olfactory bulb and nasal mucous membrane, the primary hypothesized sites for SARS-CoV-2 neuroinvasion, and the presumed source of symptoms of anosmia and ageusia reported by some infected individuals.^21^ This may be a partial explanation for the diverse neuropsychiatric symptoms^22^ displayed by many patients with severe COVID-19.

The current study reports findings from a large national survey of adults in the general population, who reported cognitive status, SARS-CoV-2 infection history, and COVID-19 symptom severity. It was hypothesized based on prior research^7,10–13^ that positive SARS-CoV-2 infection history would be associated with greater self-reported cognitive dysfunction, and that severity of COVID-19 symptoms would be positively correlated with severity of cognitive dysfunction, in a dose response manner. Finally, based on the proximity of the mOFC to the hypothesized site of neuroinvasion of SARS-CoV-2, it was expected that deficits would be evident on a delay discounting task.

## 1. Methods

### Participants

Participants were recruited as part of the Canadian COVID-19 Experiences Project (CCEP)^23^, a multi-study project which includes a national cohort survey of 1,958 adults aged 18 to 54. One research objective was to examine differences between fully vaccinated and vaccine-hesitant individuals on a broad set of demographic, psychosocial, and experiential variables. Thus, the cohort was recruited to have an equal proportion of fully vaccinated and vaccine-hesitant Canadians: 50.2% received two vaccine doses, 43.3% had received no doses, and 6.5% received one vaccine dose, but were not intending to receive a second. The mean age was 37 (*SD*=10.4) and 60.8% were female.

### Procedure

The survey was conducted from 28 September to 21 October 2021, when the predominant SARS-CoV-2 variant in Canada was Delta (4 weeks prior to the appearance of Omicron).^24^ Participants were contacted by email with an invitation to participate in the survey. A link to the survey was provided for eligible participants, and all measures were completed online following provision of informed consent. A quota target of equal number of vaccinated and vaccine hesitant was applied to obtain a balanced sample with respect to both vaccinated and vaccine-hesitant populations. Within each quota target, the sample was recruited from ten Canadian provinces through an online survey panel (Leger Opinion, the largest nationally representative probability-based panel in Canada). The survey firm and University of Waterloo monitored survey response in the sample of each quota to achieve the final representative sample. This study was reviewed and received ethics clearance from the institutional research ethics board of the University of Waterloo.

### Measures

#### Executive dysfunction

Symptoms of executive dysfunction were assessed using four “self-restraint” subscale items from the Deficits in Executive Functioning Scale, short form (BDEFS-SF)^25^. Respondents were asked how often they have experienced each the four problems during the past 6 months, including “I am unable to inhibit my reactions or responses to events or to other people”, “I make impulsive comments to others”, “I am likely to do things without considering the consequences for doing them”, and “I act without thinking”. Responses were indicated on a numerical scale where 1= never or rarely, 2=sometimes, 3=often, and 4=very often. Cronbach’s alpha for the 4 items was 0.89, indicating acceptable reliability. The four executive dysfunction items were averaged for this analysis to create a composite executive dysfunction measure.

#### Delay discounting

To assess delay discounting, participants competed a validated 5-trial delay discounting task wherein they were presented with a series of hypothetical choices between a smaller monetary amount ($500) immediately or a larger amount ($1,000) at various time delays (e.g.,1 month, 3 months).^26^ Delay discounting was calculated as a *k* value, reflecting the steepness of a hyperbolic devaluation of delayed rewards; higher values of *k* indicate more impulsive choice.

#### SARS-CoV-2 infection status

Infection status was assessed using the question “What best describes YOUR experience with [SARS-CoV-2] infection?” where 1= I have NOT been infected, 2 =I have been infected, and 3= not stated.

#### Symptom severity

COVID-19 symptom severity was assessed among those who have been infected by SARS-CoV-2 using two questions. (1) “How do you know that you HAVE BEEN infected with [SARS-CoV-2]?” responses were given the answers of 1= had symptoms but did not get tested, 2= had symptoms and tested positive, and 3 = had no symptoms but tested positive. (2) “How severe was your [SARS-CoV-2] illness?” The five-point response scale was 1=not at all severe, 2=slightly severe, 3=moderately severe, 4=very severe, 5=extremely severe. Those reporting “had no symptoms but tested positive” were incorporated into the second question as 1=not at all severe.

### Statistical analysis

Samples were post-stratified by geographic/language regions: Alberta, British Columbia, Manitoba + Saskatchewan, Ontario, Quebec English, and Quebec French, and Atlantic provinces (Nova Scotia, New Brunswick, Prince Edward Island, Newfoundland and Labrador). For each of the vaccinated and vaccine hesitant group separately, sampling weights were computed using a raking procedure and calibrated to target marginal joint population distributions of the geographic/language regions, and the gender and age group combinations, based on population figures in the 2016 Canadian census data and the disposition code in the sample, thus allowing generalization to the Canadian population. Survey linear regression models incorporating survey strata and weights were applied to estimate composite executive dysfunction scores and their associations with SARS-CoV-2 infection status and COVID-19 symptom severity. Regression models controlled for respondents’ gender and age groups (18-24, 25-39 and 40-54). All models were conducted in SAS with SUDAAN V11. All confidence intervals (CI) and statistical significance were assessed at the 95% confidence level.

## 2. Results

Baseline characteristics of the sample are presented in Table 1. The majority of the participants were female (60%) and from the 25-39 (40%) or 40-54 (43%) age groups. 84% of participants reported that they had not been infected; those who reported having been infected reported symptoms to be “not at all severe” (3%), “slightly severe” (2.4%), “moderately severe” (2.7%), with relatively few experiencing “very/extremely severe” symptoms (1%). The two cognitive measures were positively correlated (*r*=0.17, *p*<.001).

**Table 1:** Sample characteristics.

| **Variables** | ***n*** | **%** | **Executive**<br>**function**<br>**(unadjusted)**<br>**Mean, 95% CI** | **Executive**<br>**function**<br>**(adjusted)**<br>**Mean, 95% CI** |
| --- | --- | --- | --- | --- |
| Gender |  |  |  |  |
| Male | 747 | 39.27 | - | - |
| Female | 1155 | 60.73 | - | - |
| Age Group |  |  |  |  |
| 18-24 | 313 | 16.46 | - | - |
| 25-39 | 769 | 40.43 | - | - |
| 40-54 | 820 | 43.11 | - | - |
| Infection Status |  |  |  |  |
| Not infected | 1599 | 84.07 | 1.62 (1.58, 1.66) | 1.62 (1.58, 1.66) |
| Infected: Not at all severe | 57 | 3.00 | 1.72 (1.52, 1.93) | 1.73 (1.54, 1.91) |
| Infected: Slightly severe | 46 | 2.42 | 1.78 (1.44, 2.11) | 1.75 (1.45, 2.05) |
| Infected: Moderately<br>severe | 51 | 2.68 | 1.83 (1.60, 2.06) | 1.85 (1.63, 2.08) |
| Infected: Very/extremely<br>severe | 21 | 1.10 | 2.29 (1.82, 2.76) | 2.32 (1.85, 2.78) |
| Not stated | 128 | 6.73 | 1.64 (1.46, 1.81) | 1.63 (1.47, 1.80) |

Note: Executive dysfunction mean is the average of the four BDEFS items. Participants who had no COVID-19 symptoms, but tested positive for SARS-CoV-2, were classified as “not at all severe”. The adjusted parameters are adjusted by sex and group. Table 1 includes the sample used in the current analysis ( $N = 1,902$ ).

### Self-reported Executive Dysfunction

Those who reported a prior SARS-CoV-2 infection reported a significantly higher number of symptoms of executive dysfunction (*M*_*adj*_=1.89, *SE*=0.08, *CI*: 1.74, 2.04; n=175) than their non-infected counterparts (*M*_*adj*_=1.63, *SE*=0.08, *CI*: 1.47,1.80; *n*=1,599; β=0.26, *p*=.001). Men were likely to experience more executive dysfunction than women (β= 0.15, *p*<.001); younger adults (25-39 years) were more likely to experience executive dysfunction than middle aged adults (40-54 years; β= 0.30, *p*<.001).

Among those who were infected, there was a dose-response relationship between COVID-19 symptom severity and executive dysfunction. Participants who reported “moderately severe” (*M*_adj_ = 1.85, 95% *CI* 1.63 – 2.08) and “very” or “extremely severe” (*M*_adj_ = 2.32, 95% *CI* 1.85 – 2.78) COVID-19 symptoms were significantly more likely to have higher levels of executive dysfunction compared to non-infected individuals (*M*_adj_ = 1.62, 95% *CI* 1.58 – 1.66) (Table 2). A dose-response relationship between COVID-19 symptom severity and cognitive dysfunction was evident, those with moderate (β=0.23, *CI*: 0.003-0.46) and very/extremely severe (β= 0.69, *CI*: 0.22-1.16) COVID-19 symptoms being associated with significantly greater degrees of executive dysfunction, compared to those not infected and those with asymptomatic infections (Figure 2). Removing the those who reported having been intubated (*n*=5) or hospitalized without intubation (*n*=5) did not change the findings. Likewise, following further adjustment for vaccination status, income, and geographical region, those in the very/extremely severe symptom categories continued to report significantly greater symptoms of executive dysfunction than the non-infected reference group (β=0.71, 95% *CI* 0.22 - 1.19, *p*=.004).

**Table 2:** Associations between SARS-CoV-2 infection status, COVID-19 symptom severity and BDEFS scores.

| Variables | Beta (95% CI) | *p* |
| --- | --- | --- |
| Gender |  |  |
| Male | 0.15 (0.07, 0.22) | <0.001 |
| Female | Ref | Ref |
| Age Group |  |  |
| 18-24 | 0.30 (0.19, 0.41) | <0.001 |
| 25-39 | 0.06 (-0.02, 0.14) | 0.138 |
| 40-54 | Ref | Ref |
| COVID-19 Infection Status |  |  |
| Not infected | Ref | Ref |
| Infected: Not at all severe | 0.10 (-0.09, 0.29) | 0.284 |
| Infected: Slightly severe | 0.13 (-0.17, 0.42) | 0.406 |
| Infected: Moderately severe | 0.23 (0.00, 0.46) | 0.047 |
| Infected: Very/Extremely severe | 0.69 (0.22, 1.16) | 0.004 |
| Not stated | 0.01 (-0.16, 0.18) | 0.903 |

### Delay Discounting Task Performance

Participants infected with SARS-CoV-2 displayed significantly higher delay discounting rates (*k* =1.22, *SE*=0.48, *CI*: 0.27, 2.16) than non-infected participants (*k*=0.37, *SE*=0.08, *CI*: 0.21, 0.52; β=.31, *p*=.017; Table 3). With respect to dose-response effects of symptom severity, among infected individuals, those reporting “very severe” COVID-19 symptoms demonstrated significantly higher delay discounting rates than those reporting no infection history, with the remaining severity categories falling between these two values. Discount curves for infected versus non-infected, and among severity levels ranging from asymptomatic and very severe are presented in Figure 1 panel b.

**Table 3:** Associations between SARS-CoV-2 infection status, COVID-19 symptom severity and delay discounting.

| Variables | Beta | p |
| --- | --- | --- |
| *Infection Status* |  |  |
| COVID-19 infection status |  |  |
| Infected | 0.31 (0.06, 0.56) | 0.017 |
| Not infected | Ref | Ref |
| Not stated | -0.01 (-0.22, 0.20) | 0.91 |
| *Symptom Severity* |  |  |
| Gender |  |  |
| Male | -0.10 (-0.20, 0.01) | 0.066 |
| Female | Ref | Ref |
| Age group |  |  |
| 18-24 | -0.04 (-0.18, 0.11) | 0.618 |
| 25-39 | 0.01 (-0.11, 0.13) | 0.893 |
| 40-54 | Ref | Ref |
| Income |  |  |
| Low | Ref | Ref |
| Moderate | 0.02 (-0.17, 0.21) | 0.838 |
| High | -0.30 (-0.45, -0.16) | <0.001 |
| No answer | -0.34 (-0.57, -0.12) | 0.002 |
| COVID-19 infection status |  |  |
| Not infected | Ref | Ref |
| Infected: Asymptomatic | -0.01 (-0.29, 0.27) | 0.934 |
| Infected: Slightly severe | 0.24 (-0.09, 0.57) | 0.147 |
| Infected: Moderately severe | 0.34 (-0.11, 0.79) | 0.141 |
| Infected: Very severe | 1.26 (0.31, 2.21) | 0.009 |
| Not stated | -0.01 | 0.92 |

**Figure 1.**
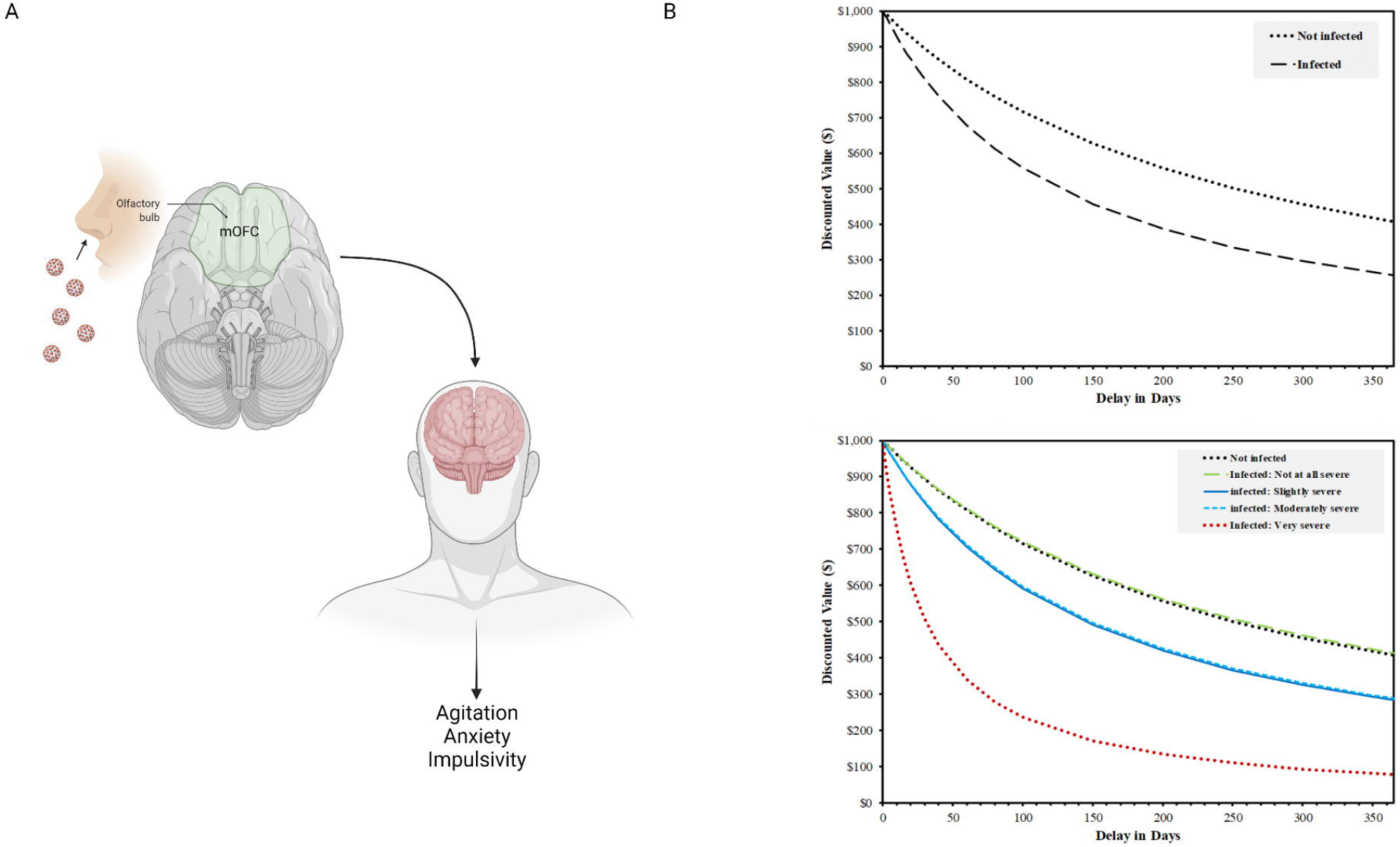
Conceptual diagram (A) and delay discounting curves for non-infected and ranges of COVID-19 symptom severity from asymptomatic to “very severe” (B).

**Figure 2.**
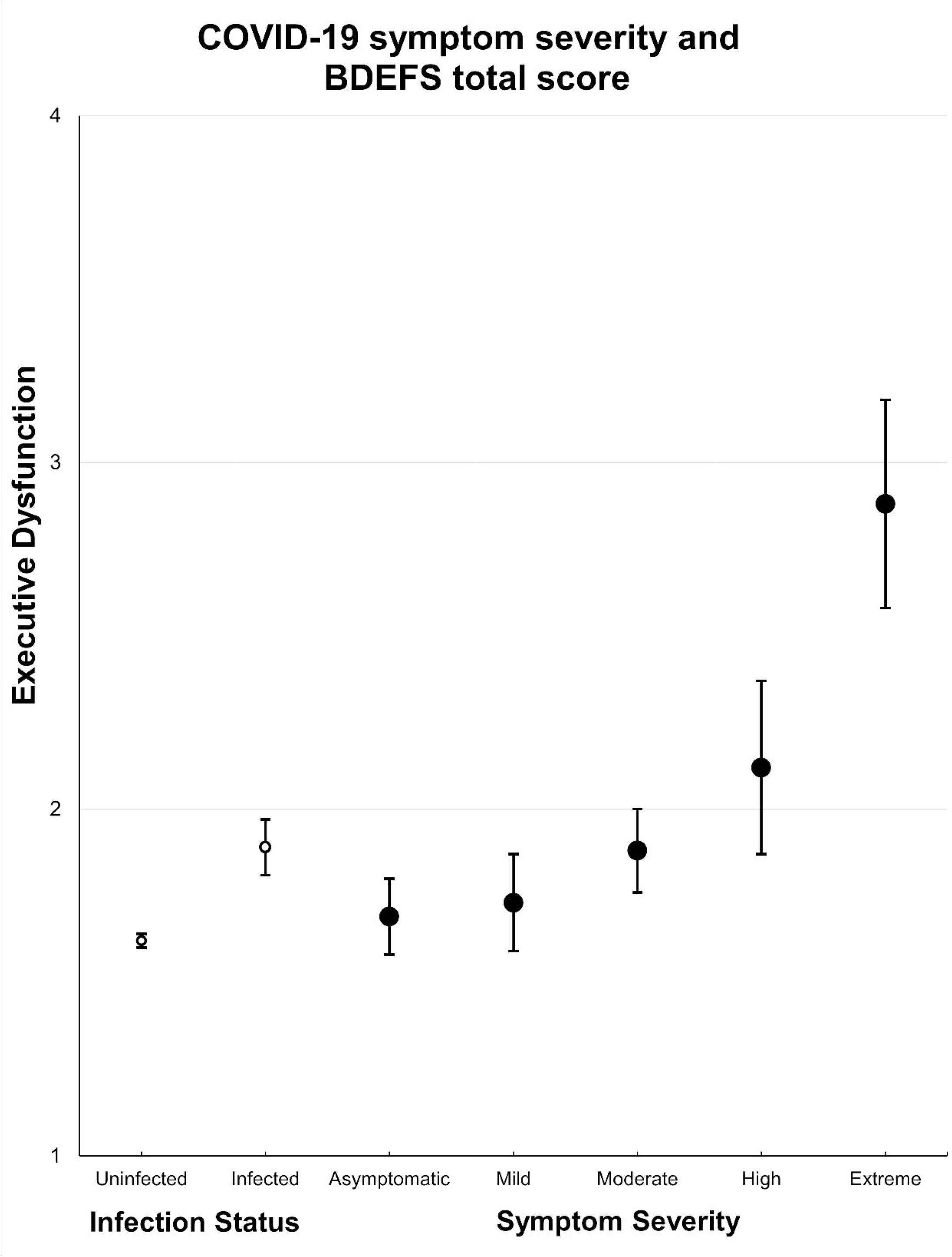
Effects of SARS-CoV-2 infection status and COVID-19 symptom severity on BDEFS scores; BDEFS=Barkley Deficits in Executive Functioning Scale.

In general, males had marginally steeper discount rates than females (β=-.10, *p*=.066), and individuals reporting high incomes had significantly lower discounting rates than individuals reporting low income (β=-.30, *p*<.001). No significant age differences in k values were observed (see supplementary materials). No two-way interactions were observed between sex and infection status predicting delay discounting were observed (Wald *F*=0.09, *p*=0.91), or between age and infection status predicting delay discounting (Wald *F*=0.90, *p*=0.46). Likewise, the three-way interaction term between sex, age and infection status in predicting delay discounting was non-significant (Wald *F*=1.37, *p*=0.22).

### Sensitivity Analyses

Further adjustment for education and geographical region (i.e., province) had no overall effect on the findings. In education and province-adjusted models, those reporting a SARS-CoV-2 infection continued to show a significantly greater degree of delay discounting than those non-infected (β=-0.32, *CI*:-0.57,-0.06, *p*=.014). Also similar to earlier analyses, those in the “very severe” COVID-19 symptom severity category showed greater discounting than those in the non-infected group (β=1.28, *CI*: 0.35,2.21, *p*=.007). Likewise, removal of 5 cases reporting being placed on mechanical ventilator did not change the effects of SARS-CoV-2 infection status (β=.23, *CI*: 0.01,0.45, *p*=.043) or COVID-19 symptom severity (β=.95, *CI*: 0.20,1.71, *p*=.014) on delay discounting rate. Finally, when limiting the “infected” group to only those whom reported having their infection confirmed by a positive PCR test, the effect of SARS-CoV-2 infection remained significant, and somewhat stronger in magnitude (β=.40; CI: 0.07, 0.72, *p*=.016).

## 3. Discussion

In this population-representative cohort of community-dwelling adults, those with a positive history of SARS-CoV-2 infection reported more symptoms of cognitive dysfunction than those with no such history. This effect was evident on both self-reported symptoms of executive dysfunction and on a validated decision-making task. A dose-response relationship between COVID-19 symptom severity and magnitude of cognitive dysfunction was evident such that increasing infection severity was associated with greater symptoms of cognitive dysfunction for both self-reported symptoms and task performance. Importantly, reliable effects of positive SARS-CoV-2 infection history and COVID-19 symptom severity on cognitive dysfunction were evident—on both measures—even in this sample of individuals not typically subject to age-related cognitive decline (ages 18 to 54) and not exposed to medically induced coma via hospital-based treatment for severe COVID-19. Our findings were similar to a prior report of executive dysfunction as correlated with COVID-19 symptom severity in a large population sample^13^, but extend them to include self-reported symptoms of interpersonal significance, and a standardized decision making paradigm previously linked to the site of hypothesized neuroinvasion of the SARS-CoV-2 virus (the mOFC; Figure 1 panel A).

There are several hypothesized mechanisms by which SARS-CoV-2 infection may produce cognitive dysfunction, including encephalitis, coagulopathy, cytokine storm, hypoxia, and megakaryocyte invasion.^4–6^ The current investigation cannot distinguish among these neurophysiological mechanisms, or others that may yet be identified. The current findings do not preclude the possibility that symptoms of cognitive dysfunction are influenced by reporting biases among those who are continuing to experience emotional distress following the measurement period. Given that the effects of negative mood on symptom reporting is causally established,^27,28^ and given that mood impacts of the COVID-19 pandemic are well-documented,^29–33^ this possibility cannot be definitively excluded. However, at least one prior population-based study has found similar dose-response effects using performance-based measures of cognitive function (i.e., cognitive tasks rather than reported symptoms).^7^ It is further noteworthy that the same patterns were evident on our decision-making task.

It is not clear why there appeared to be a stronger link between SARS-CoV-2 infection and cognitive dysfunction in younger adults as compared with middle-aged adults. It is possible that such deficits were more salient to younger adults, given that a higher proportion would be in educational programs wherein lapses in attention and concentration may have been more impactful. In either case, it is not clear how consequential symptoms of cognitive dysfunction would be expected to be, even if reliable across studies. It is not uncommon for other types of viral infections to cause symptoms of cognitive dysfunction, including the seasonal flu, herpes, MERS, Zika and Varicella (chickenpox).^34–38^ Documenting the stability and functional impact of any SARS-CoV-2 infection impairments in cognition will be important. However, in the meantime, reductions in unnecessary exposure to SARS-CoV-2 infection may be an important public health strategy even for young and middle aged adults, despite the limited mortality risk.

Finally, given that the predominant SARS-CoV-2 variant during the time of the survey was Delta, the findings are applicable only to the Delta and earlier variants. Moreover, the retrospective nature of the study does not allow us to determine with confidence which infections were attributable to Delta versus earlier variants. We also cannot conclude that the same associations would be observed with the Omicron variant, in particular because of the lower COVID-19 symptom severity apparent with Omicron in comparison with earlier variants, at least based on early data.^39–41^ In the current (pre-Omicron) sample, we found that only moderate and higher COVID-19 symptom severities were associated with significantly elevated symptoms of executive dysfunction. Further analyses of follow-up waves of the CCEP data will enable examination of the relative impact of the Omicron variant on symptoms of executive dysfunction.

### Strengths and Limitations

There are several strengths of the current study. One strength is the use of a large population-representative sample, consisting of infected individuals of a wide range of disease symptom severities—ranging from asymptomatic to hospitalized—as well as non-infected controls. Another strength is the use of a validated measure of subjective symptomology assessing everyday function rather than more sensitive but less ecologically valid performance-based measures. Finally, the finding of similar effects on a decision-making task performance increases confidence that the findings were not a function of self-report methodology alone. In terms of limitations, by virtue of the survey format, it was not possible to validate the infection status of individuals by testing. This may lead to under-or over-estimation of effect size and statistical significance of tests, vis-a-vis misreporting of infection status. This is a limitation of many survey studies of COVID-19 and cognitive dysfunction, however. Finally, the cross-sectional design limits our ability to draw causal inferences.

Future studies should examine the longevity of cognitive dysfunction symptoms over time, as well as the extent to which the dose-response and age gradients observed here are replicable across samples. Finally, additional studies examining neurological impacts at the level of the brain itself will be required, using functional brain imaging paradigms to quantify structural and functional impacts of SARS-CoV-2 infection. In particular studies are needed that follow individuals forward from the point of infection to examine changes over time, in a prospective manner.

## Conclusions

In summary, the current study used a population-representative sample consisting of a balanced proportion of vaccinated and unvaccinated individuals to estimate the association between SARS-CoV-2 infection and symptoms of cognitive dysfunction. Findings indicated that individuals previously infected with SARS-CoV-2 reported significantly greater symptoms of cognitive dysfunction than non-infected individuals. Further, among those reporting a positive infection history, a dose-response relationship between COVID-19 symptom severity and cognitive dysfunction was evident, such that those with moderate to severe symptoms were more likely to experience symptoms of cognitive dysfunction. The above pattern was evident for both self-reported symptoms of cognitive dysfunction and performance on a decision-making task. Taken together with findings from other studies, cognitive dysfunction appears to be a correlated of SARS-CoV-2 infection, particularly among those with at least moderate COVID-19 symptom severity. If such cognitive effects are long-lasting, this may be one piece of evidence in support of public health strategies that eliminate exposure to SARS-CoV-2 infection, even for young adults and those below the typical high-risk age threshold for mortality.

## Data Availability

All data produced in the present study are available upon reasonable request to the authors

## Research ethics statement

This study protocol was reviewed by and received approval from the University of Waterloo Office of Research Ethics.

## Data Availability Statement

Data will be available upon reasonable request to either of the corresponding authors.

## Acknowledgements

We thank Anne C.K. Quah and Thomas Agar for their assistance with survey design and management.

## Author Contributions

PH, GF, and SH conceived the study, planned and oversaw the statistical analyses, and wrote the final draft. GM planned and completed all statistical analyses and contributed to the writing of the final draft. MNS, AH, JM, and WB contributed to the writing of the final draft.

